# Returning *APOE* and pTau-217 Results: the eSMARTER Randomized Noninferiority Clinical Trial

**DOI:** 10.64898/2026.08.27.26361535

**Authors:** Jessica B. Langbaum, Claire M. Erickson, Carolyn Langlois, Elisabeth M. Wood, Brian L. Egleston, Kristin Harkins, Rajia Mim, Samantha John, Camille Brown, Sarah Brown, Sarah Howe, Cara Cacioppo, Laura Eppelmann, Jessica Enos, Hayley Salata, Diana DeSantiago, Emily A. Largent, Eric M. Reiman, Marisa N. Denkinger, Nicholas J. Ashton, J. Scott Roberts, Jason Karlawish, Angela R. Bradbury

**Affiliations:** Banner Alzheimer’s Institute, Phoenix, Arizona; Department of Medicine, Division of Hematology-Oncology, University of Pennsylvania Perelman School of Medicine, Philadelphia, Pennsylvania; Fox Chase Cancer Center, Temple University Health System, Philadelphia, Pennsylvania; Department of Medicine, Division of Geriatrics, University of Pennsylvania Perelman School of Medicine, Philadelphia, Pennsylvania; Department of Medical Ethics and Health Policy, University of Pennsylvania Perelman School of Medicine, Philadelphia, Pennsylvania; Banner Sun Health Research Institute, Sun City, Arizona; Department of Health Behavior and Health Equity, University of Michigan School of Public Health, Ann Arbor, Michigan; Dignity Health St Joseph’s Hospital and Medical Center, Phoenix, Arizona

## Abstract

**Importance:** Patients are increasingly learning Alzheimer’s disease (AD) genetic and biomarker results through electronic health portals. Evaluation of alternative scalable delivery models for return of AD risk information is needed to best support patient understanding and psychological well-being.

**Objective:** To determine whether a patient-centered digital platform is comparable to clinician-mediated telehealth sessions for returning *APOE* and plasma pTau-217 results on outcomes of knowledge and psychological well-being.

**Design:** The Evaluation of Self-Mediated Alternatives for Risk Testing Education and Return of Results (eSMARTER) study was a noninferiority trial of a patient-centered digital platform compared to clinician-mediated disclosure of *APOE* genotype and optional pTau-217 disclosure.

**Setting:** Decentralized, fully remote trial enrolled participants in the contiguous United States (U.S.) between October 2024 and February 2025, with follow-up completed in November 2025.

**Participants:** Eligible participants were aged 60-80 and had previously undergone *APOE* genotyping (without disclosure) via the GeneMatch program, passed psychological screening, had internet access, and were English-speaking.

**Interventions:** Participants were randomized, 2:1, to the eSMARTER digital platform or clinician-mediated disclosure of *APOE* genotype. Following the 6-month post-*APOE* assessment, participants were offered optional pTau-217 disclosure via the same randomized modality.

**Main Outcomes and Measures:** Primary outcomes at 1-7 days following *APOE* disclosure included changes in anxiety, disease-specific distress, and AD-related knowledge within a priori non-inferiority margins.

**Results:** 674 persons (mean [SD] age 68 [4.7] years; 451 [67%] female; mean [SD] telephone MoCA=19 [2]) were eligible and provided demographic information. 651 participants were randomized to clinician-mediated (n=216) or digital disclosure (n=435) and completed *APOE* disclosure (66 [10%] *APOE4* homozygotes, 377 [58%] heterozygotes, 208 [32%] non-carriers). 604 participants completed the study; 500 completed optional pTau-217 disclosure. Baseline characteristics were balanced across groups. At 1-7 days following *APOE* disclosure, scores on AD-related knowledge, PROMIS Anxiety, and disease-specific distress measures met non-inferiority.

**Conclusions and Relevance:** Disclosure of *APOE* genotype by the eSMARTER digital platform is non-inferior to clinician-mediated telehealth disclosure. No significant between group differences were found following disclosure of pTau-217 results. Together, these results suggest that this digital platform may provide an evidence-based scalable approach for returning AD genetic and biomarker results.

**Trial Registration:** ClinicalTrials.gov Identifier NCT06459583

## Introduction

The 21^st^ Century Cures Act mandates the immediate return of most medical test results to patients in the United States (U.S.), including Alzheimer’s disease (AD) genetic and biomarker results.^1^ Consequently, patients can view test results in their electronic health portal before speaking with their healthcare provider. Although prior studies demonstrate that disclosing *APOE* genotype and brain amyloid neuroimaging results can be conducted in a generally safe and well-tolerated manner,^2–13^ these all relied on clinicians to conduct the disclosure visits, either in-person or via telehealth.^14^ This limits the clinical translatability of these prior findings, as patients are now often first learning results independently and without explanation via electronic health portals. Due to ongoing shortages of clinicians equipped to return and discuss AD-related results, opportunities for clarification and support may be limited and difficult to access. Additionally, current standard of care return of result protocols require considerable clinician time, further limiting the broader translatability of prior research investigating the impact of returning AD-related results. Scalable, evidence-based approaches to returning AD-related information are therefore needed. Yet, it remains unknown whether return of results via digital platforms, such as one that could be integrated into an electronic health portal, is comparable to a clinician-mediated telehealth session in terms of patient safety, well-being, and understanding of the information.

Digital health technologies offer an innovative and scalable pathway for communicating AD genetic and biomarker results. Such results can be helpful in identifying individuals at elevated disease risk and/or in informing early detection of AD. Given the anticipated shift in AD screening and testing from specialty memory clinics to primary care, safe and effective self-mediated digital platforms that can be scaled for widespread use are needed.^15^ These digital platforms can be developed to include structured education and personalized communication of results, allowing the patient and care partner(s) to review the information as needed. They have the potential to reduce clinician burden and patient misinterpretation of results.^16^

Based on previous findings from the Alzheimer’s Prevention Initiative (API) Generation Program’s *APOE* disclosure program^14^ and its ancillary CONNECT4 APOE randomized trial of telephone versus telehealth videoconference disclosure of *APOE* results,^17^ we designed the subsequent eSMARTER clinical trial. eSMARTER was a randomized, non-inferiority trial comparing returning *APOE* results to participants via digital platforms or a clinician telehealth session, with optional pTau-217 disclosure, a biomarker of AD pathology, 6-months after returning *APOE* results, by assessing changes in measures of anxiety, disease-specific distress, and AD-related knowledge following disclosure.^18^

## Methods

### Trial Design and Conduct

The eSMARTER trial was a decentralized and fully remote trial to assess the comparability of returning AD genetic and biomarker results from self-mediated, patient-informed digital platform or via clinician telehealth (genetic counselor [GC] for *APOE* results, physician for pTau-217 results). The study design was published previously.^18^ Enrollment was from 10-Oct-2024 to 11-Feb-2025; data collection was completed on 7-Nov-2025. The eSMARTER protocol was approved by an Advarra institutional review board (IRB) (#Pro00064250), and all participants provided written informed consent.

Participants received gift cards for survey completions and the blood draw visit. An independent blinded safety officer reviewed and oversaw study safety. Reporting followed Consolidated Standards of Reporting Trials (CONSORT) guidelines.

### Participants

Participants were recruited from GeneMatch, a trial-independent research enrollment program that performs *APOE* genotyping in labs accredited by the College of American Pathologists (CAP) and certified through Clinical Laboratory Improvement Amendments (CLIA).^19^ GeneMatch sent participation invitation emails and mailed letters to eligible members.

Inclusion criteria included: being age 60-80 at screening; having undergone prior *APOE* genetic testing via GeneMatch without having received results; having access to the internet and an internet-enabled device; speaking English; and being willing to undergo a blood draw.. Exclusion criteria were prior knowledge of *APOE* genotype, lack of psychological readiness to receive *APOE* and pTau-217 results as indicated by a score of 10 or greater on the Patient Health Questionnaire – 9 (PHQ-9)^20^ and/or responding yes to any question from the abbreviated Columbia Suicide Severity Scale (C-SSRS) or AD Specific Suicidality Questions, presence of a current untreated or unstable major psychiatric illness, and communication difficulties. Eligible participants completed the Telephone Montreal Cognitive Assessment (T-MoCA)^21^ and provided additional screening demographics.

### Digital interventions for pre-disclosure education and return of results

The trial compared two methods (patient-informed digital platform or clinician telehealth) of returning *APOE* genotype and associated risk of mild cognitive impairment (MCI) or dementia by age 85 using risk estimates calculated for the API Generation Program.^22,23^ These methods were used to disclose categorical pTau-217 results (negative, intermediate, positive).^18^ The eSMARTER scalable digital platforms (*APOE*, pTau-217) are informed by a modified tiered-binned model and user tested by age-matched participants.^18,24^ Content and modules have been previously described and were designed to cover what a GC or clinician would address in pre-test education and disclosure of results.^18^ Tier 1 information included indispensable information presented to all users; Tier 2 contained optional, more in-depth content. Print and electronic materials containing all content were developed for the clinicians who were trained prior to conducting telehealth sessions. To better understand differences in user preferences for different platforms, we developed web-based and SMS-based chat platforms for *APOE* and pTau-217 disclosure. Participants in the digital arm selected which platform (web or chat) they preferred. All participants randomized to the digital arm could request to speak with a clinician before, after or instead of using the digital platform. All participants were offered optional digital educational materials about *APOE* and pTau-217 to review prior to the disclosure sessions. Procedures for both arms are previously described.^18^

### Randomization

After screening assessments, participants were randomized 2:1 to digital platforms or clinician telehealth disclosure. Randomization was by permuted block design and performed centrally using FlexAdvantage with stratification factors of sex (male, female, other), *APOE4* carrier group (non-carrier, heterozygote, homozygote). Study personnel did not have access to the allocation sequence. *APOE* genotype results used for disclosure were provided by GeneMatch.^19^

### Outcomes

Outcomes to evaluate cognitive, affective and behavioral outcomes of digital vs telehealth disclosure were collected at baseline (T0), 1-7 days (T1), 6 weeks (T2) and 6 months (T3) after *APOE* disclosure, and 1-7 days (T4) and 4 weeks (T5) after optional pTau-217 disclosure (**Supplemental eTable1**). Remote phlebotomy services were used to collect blood between visits T2 and T3; pTau-217 concentrations were measured on the Roche cobas e801 platform using a Research Use Only (RUO) assay to provide categorical results.^18^

The primary outcomes of *APOE* disclosure included: 1) AD-related knowledge (with focus on *APOE*) assessed by 8 items adapted from the Cancer Genetics and ClinSeq Knowledge Scales (Cronbach α = 0.83);^25–27^ 2) Anxiety assessed by the 7-item Patient Reported Outcomes Measurement Information System (PROMIS) Anxiety measure (Cronbach α = 0.79);^28,29^ and 3) Disease-specific distress measured by 8 items adapted from the Impact of Events Scale.^30,31^

Secondary outcomes of *APOE* disclosure included Patient Health Questionnaire-9 (PHQ-9),^20^ satisfaction with genetic services (single item),^32–34^ Psychological Wellbeing Scale,^35^ perceived risk of AD,^36,37^ impact of genetic testing for AD (IGT-AD),^38^ recall of results, Cognitive Function Instrument,^39,40^ and modified Stigma Impact Scale.^41^ These outcomes were also assessed for pTau-217 disclosure and considered secondary.

### Covariates

Age, sex, race (available choices from which participants could select included American Indian or Alaskan Native, Asian, Black or African American, Caucasian or White, Hawaiian or other Pacific Islander, Other, Unknown; more than one race could be selected), ethnicity (Hispanic or Latino, not Hispanic or Latino), highest degree or level of school completed, family history of AD, marital status, employment status, and household income were collected via self-report. Collection of race and ethnicity was required by the funding agency. Other covariates included Area Deprivation Index (ADI) score and rurality, calculated using 2019 National Center for Education Statistics.42

## Statistical Analysis

We used t-tests and Chi-squared tests to compare demographic differences between arms. We hypothesized that digital platform disclosure of *APOE* would result in non-inferior outcomes compared to clinician-mediated (telehealth) usual care. For primary analyses, we would conclude non-inferiority if the upper bound of the randomization effect 96.7% confidence interval (CI) for each behavioral change variable did not cross the non-inferiority threshold in a direction that favors usual care. The non-inferiority margins were 2 points for genetic knowledge, 3.5 points for anxiety, and 0.35 baseline standard deviations for disease-specific distress. We used clinical judgment to set the margin for AD-related knowledge. We used the midpoint between a small (0.2 standard deviation [SD]) and large (0.5 SD) effect for disease-specific distress (0.35 SD is midpoint). We set the margin for PROMIS anxiety t-score 0.5 points below the value of 4 that has been reported as clinically relevant.^43^ Based on preliminary data, we had 85% non-inferiority power and 1.67% Type I error (1-sided). We chose 1.67% Type I error by applying a Bonferroni correction to a 5%. We used multiple imputation methods for the outcome analyses.^44^ In secondary analyses, we tested for differences among arms using t-tests, including for *APOE4* carrier subgroups (homozygotes and heterozygotes combined versus non-carriers). For secondary analyses, we set the threshold for statistical significance to p<0.01.

## Results

### Participants

Enrollment, randomization, and survey completion are shown in **Figure 1**. Of the 813 participants who consented, 681 were eligible for randomization, 666 completed the baseline survey, 651 were randomized and received *APOE* disclosure. Of these, 600 (90%) completed optional digital education prior to disclosure. The mean (SD) age of participants was 67.9 (4.7) years, 439 (67%) women, 585 (90%) White, 38 (6%) Hispanic, and 143 (22%) had less than a college degree (**Table 1**). Participants were enrolled from the contiguous U.S., 52% were from rural areas. Mean T-MoCA score was 19.0 (SD 2.0) and 232 (36%) participants had scores <19, a cutoff commonly used for cognitive impairment. 434 (67%) participants had a family history of AD; 66 (10%) were *APOE4* homozygotes, 377 (58%) heterozygotes, and 208 (32%) non-carriers. 126 (22%) of participants had a positive pTau-217 result, 175 (31%) had an intermediate result, and 264 (47%) had a negative result. There were no significant differences in baseline characteristics between arms.

**Figure 1.**
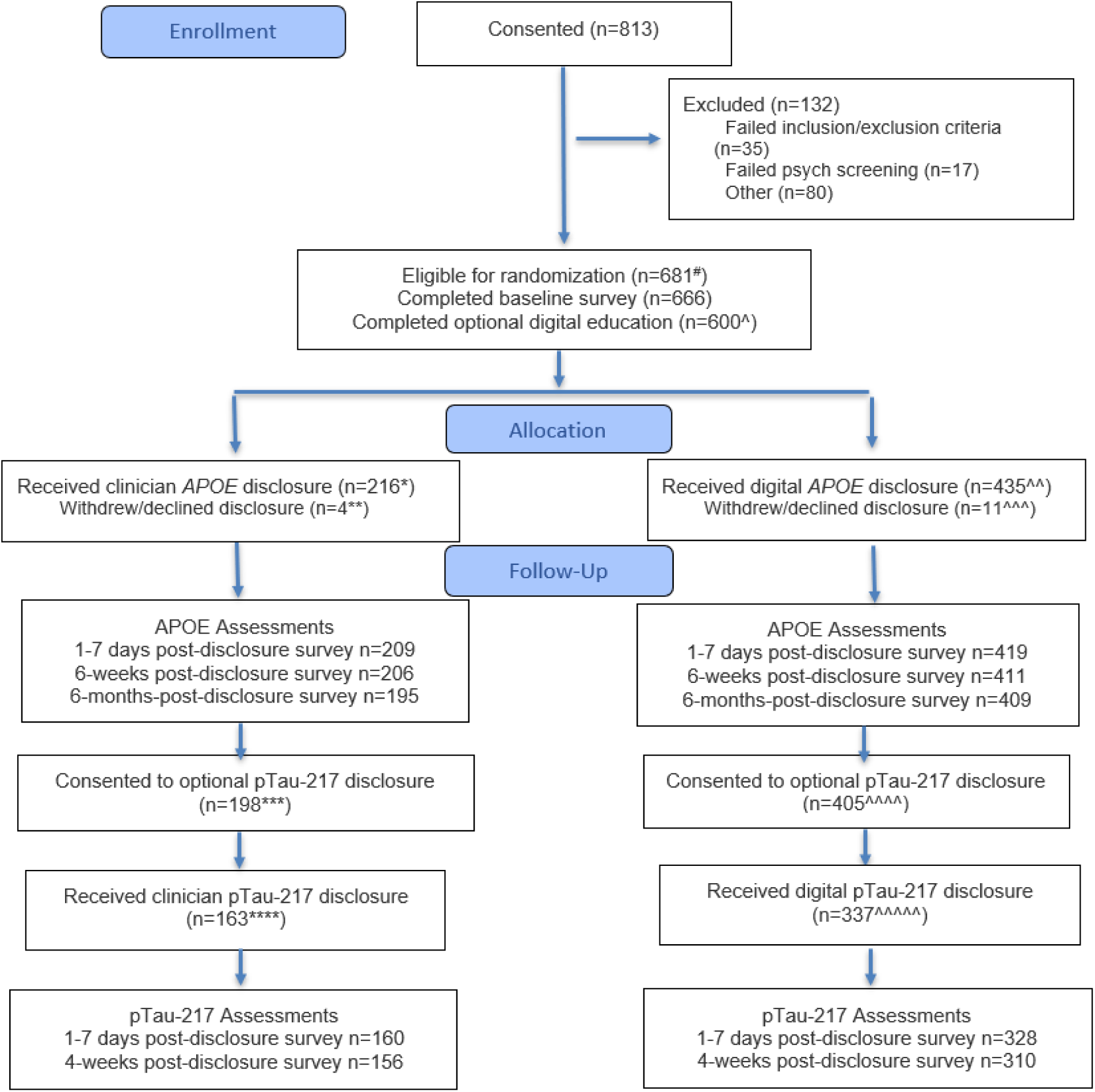
CONSORT Diagram. eSMARTER study profile illustrating eligibility screening before the Baseline Survey, after completion of the Baseline Survey, and after Randomization. Randomization was performed on participants selected for the trial who completed the Baseline Survey and was stratified by sex (male, female, other), *APOE4* carrier group (non-carrier, heterozygote, homozygote).

**Table 1.** Sample Characteristics by Randomization Arm.

|  | Randomization Arm |  |  | p-value |
| --- | --- | --- | --- | --- |
|  | Telehealth<br>(N = 216) | Digital<br>(N = 435) | Total<br>(N = 651) |  |
| **Age** | 68.1 (4.8) | 67.8 (4.7) | 67.9 (4.7) | 0.39 |
| **Sex** |  |  |  | 0.81 |
| Female | 147 (68%) | 292 (67%) | 439 (67%) |  |
| Male | 69 (32%) | 143 (33%) | 212 (33%) |  |
| **Race** |  |  |  | 0.35 |
| American Indian or Alaska Native | 1 (1%) | 3 (1%) | 4 (1%) |  |
| Asian | 3 (1%) | 10 (2%) | 13 (2%) |  |
| Black or African American | 8 (4%) | 13 (3%) | 21 (3%) |  |
| White | 198 (92%) | 387 (89%) | 585 (90%) |  |
| Other | 6 (3%) | 13 (3%) | 19 (3%) |  |
| Not known | 0 (0%) | 9 (2%) | 9 (1%) |  |
| **Ethnicity** |  |  |  | 0.89 |
| Not Hispanic | 203 (94%) | 410 (94%) | 613 (94%) |  |
| Hispanic | 13 (6%) | 25 (6%) | 38 (6%) |  |
| **Marital Status** |  |  |  | 0.92 |
| Currently married or partnered | 156 (72%) | 312 (72%) | 468 (72%) |  |
| Widowed | 17 (8%) | 40 (9%) | 57 (9%) |  |
| Divorced or separated | 32 (15%) | 63 (15%) | 95 (15%) |  |
| Never married | 11 (5%) | 19 (4%) | 30 (5%) |  |
| Unknown | 0 (0%) | 1 (0%) | 1 (0%) |  |
| **Education Level** |  |  |  | 0.85 |
| High School or less | 8 (4%) | 16 (4%) | 24 (4%) |  |
| Some College or trade school | 37 (17%) | 82 (19%) | 119 (18%) |  |
| College or more | 171 (79%) | 336 (77%) | 507 (78%) |  |
| Unknown | 0 (0%) | 1 (0%) | 1 (0%) |  |
| **Family History of Alzheimer's** |  |  |  | 0.20 |
| No | 46 (21%) | 83 (19%) | 129 (20%) |  |
| Yes | 148 (69%) | 286 (66%) | 434 (67%) |  |
| Not sure | 22 (10%) | 66 (15%) | 88 (14%) |  |
| **Telephone Montreal Cognitive Assessment (MoCA) Total Score (0-22)** | 19.1 (1.9) | 19.0 (2.1) | 19.0 (2.0) | 0.70 |
| **Health Literacy at Baseline (3-15)** | 7.06 (1.02) | 6.93 (1.07) | 6.97 (1.05) | 0.13 |
| **Area Deprivation Index Score (1-100)** | 36.7 (24.0) | 39.4 (23.7) | 38.5 (23.8) | 0.17 |
| **Rurality** |  |  |  | 0.94 |
| City/Suburb/Town | 101 (47%) | 207 (48%) | 308 (37%) |  |
| Rural | 113 (52%) | 223 (51%) | 336 (52%) |  |
| Unknown | 2 (1%) | 5 (1%) | 7 (1%) |  |

|  | Randomization Arm |  |  |  |
| --- | --- | --- | --- | --- |
|  | Telehealth<br>(N = 216) | Digital<br>(N = 435) | Total<br>(N = 651) | p-value |
| **Income** |  |  |  | 0.19 |
| <\$100,000 | 107 (50%) | 222 (51%) | 329 (50%) | |
| ≥\$100,000 | 99 (46%) | 178 (41%) | 277 (43%) | |
| Prefer not to answer | 10 (5%) | 35 (8%) | 45 (7%) |  |
| **Employment Status** |  |  |  | 0.16 |
| Employed/Homemaker | 68 (32%) | 139 (32%) | 207 (32%) |  |
| Not working | 12 (6%) | 10 (2%) | 22 (3%) |  |
| Retired | 136 (63%) | 285 (66%) | 421 (65%) |  |
| Unknown | 0 (0%) | 1 (0%) | 1 (0%) |  |
| **APOE Genotype** |  |  |  | 0.98 |
| Homozygote | 22 (10%) | 44 (10%) | 66 (10%) |  |
| Heterozygote | 124 (57%) | 253 (58%) | 377 (58%) |  |
| Non-carrier | 70 (32%) | 138 (32%) | 208 (32%) |  |
| **pTau-217 Results by APOE Carriership** |  |  |  | 0.16 |
| Positive pTau-217 |  |  |  |  |
| Carrier | 41 (19%) | 68 (16%) | 109 (17%) |  |
| Non-Carrier | 10 (5%) | 7 (2%) | 17 (3%) |  |
| Intermediate pTau-217 |  |  |  |  |
| Carrier | 41 (19%) | 89 (21%) | 130 (20%) |  |
| Non-Carrier | 10 (5%) | 35 (8%) | 45 (7%) |  |
| Negative pTau-217 |  |  |  |  |
| Carrier | 46 (21%) | 102 (23%) | 148 (23%) |  |
| Non-Carrier | 37 (17%) | 79 (18%) | 116 (18%) |  |
| Unknown (i.e., not tested) | 31 (14%) | 55 (13%) | 86 (13%) |  |

Abbreviation: ADI, Area Deprivation Index; APOE, apolipoprotein E; MoCA, Montreal Cognitive Assessment

Failure to have *APOE* disclosure, and hence exclusion from ITT analyses, did not differ between arms (8 of 224 [4%] initially randomized to telehealth; 15 of 450 [3%] initially randomized to digital) (**Supplemental eTable 2**). Participants who did not receive results were older (69.9 [SD 4.7] years, not disclosed, versus 67.9 [SD 4.7] years disclosed; *P*=0.048), and more likely to be Hispanic (17% of those not disclosed versus 6% disclosed; *P*=0.024).

### Primary Non-inferiority analyses for *APOE* disclosure

In the primary intention-to-treat (ITT) analyses, we met the non-inferiority thresholds for all primary outcomes (**Figure 2**). We also met the non-inferiority thresholds for all secondary outcomes and all timepoints (T0-T3). Two (0.5%) participants randomized to the digital arm requested a GC. Per-protocol non-inferiority analyses were similar to the ITT results.

**Figure 2.**
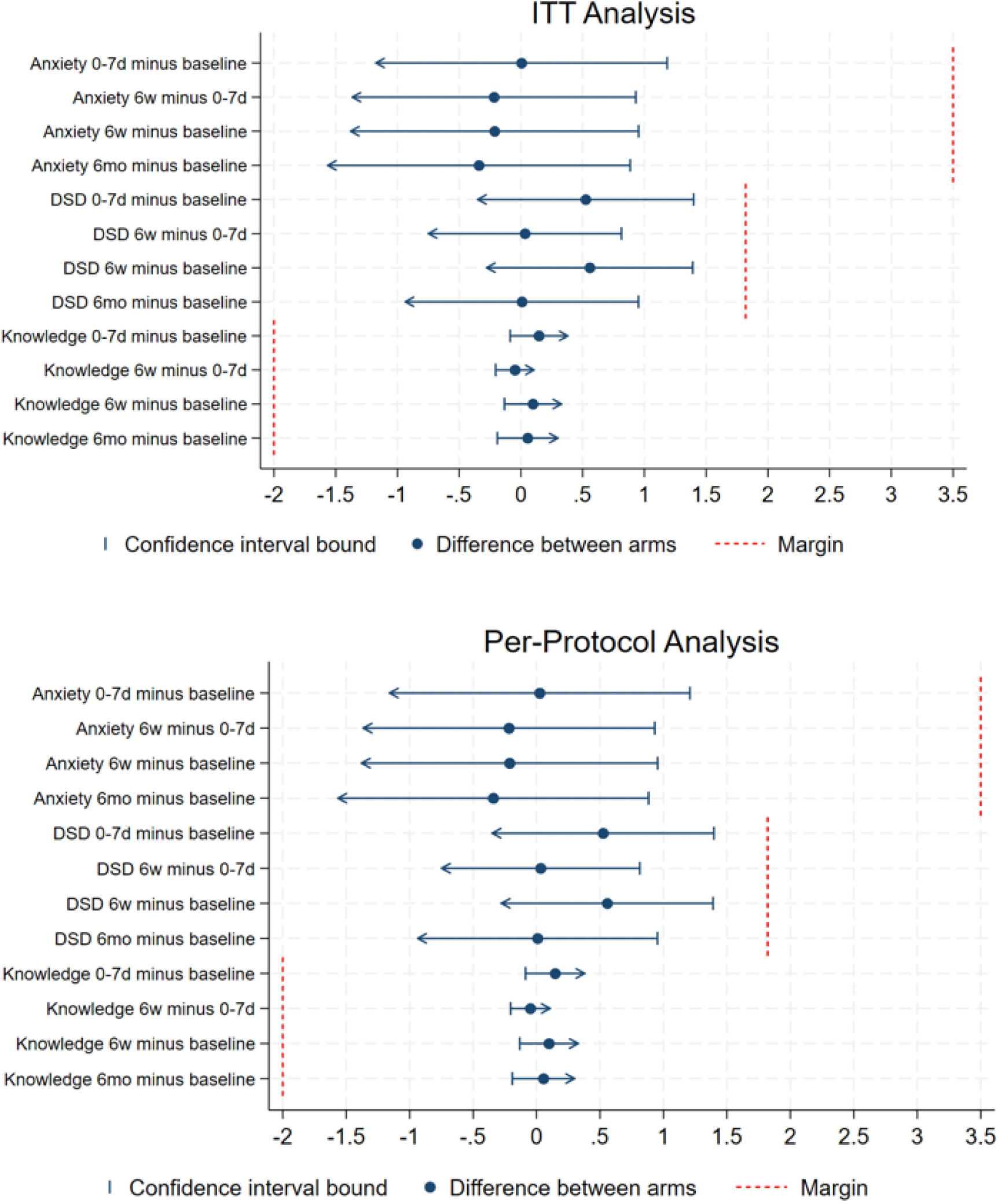
Non-inferiority Intention-to-Treat (ITT) and per-protocol analyses. The blue dots represent the difference between arms by each primary outcome: change in anxiety, change in disease specific distress (DSD), and change in APOE knowledge. The vertical dotted line represents the non-inferiority margin. Failure to cross the margin threshold means digital disclosure was non-inferior to telehealth disclosure on the relevant item.

### Secondary analyses for *APOE* disclosure

As shown in **Table 2**, most primary and secondary outcomes did not differ significantly by arm in the ITT analyses. There was slightly higher negative response on the IGT-AD in the digital compared to the telehealth arm immediately post-disclosure (T1) (14.1 [SD 11.6] vs 11.3 [SD 9.8]; *P*=0.0026), although there were no differences at later time points and the magnitude of the difference is small, suggesting these may not be clinically significant. Satisfaction was overall high, but higher in the telehealth vs digital arm (4.8 [SD 0.7] vs 4.3 [SD 0.9]; *P* < 0.001). There was slightly lower accuracy in recall of *APOE* result at T2 in the digital compared to the telehealth arm (14% incorrect vs 6% incorrect; *P* = 0.0054), although there were no differences at T1 or T3 timepoints. These findings were similar in per-protocol analyses.

**Table 2.**
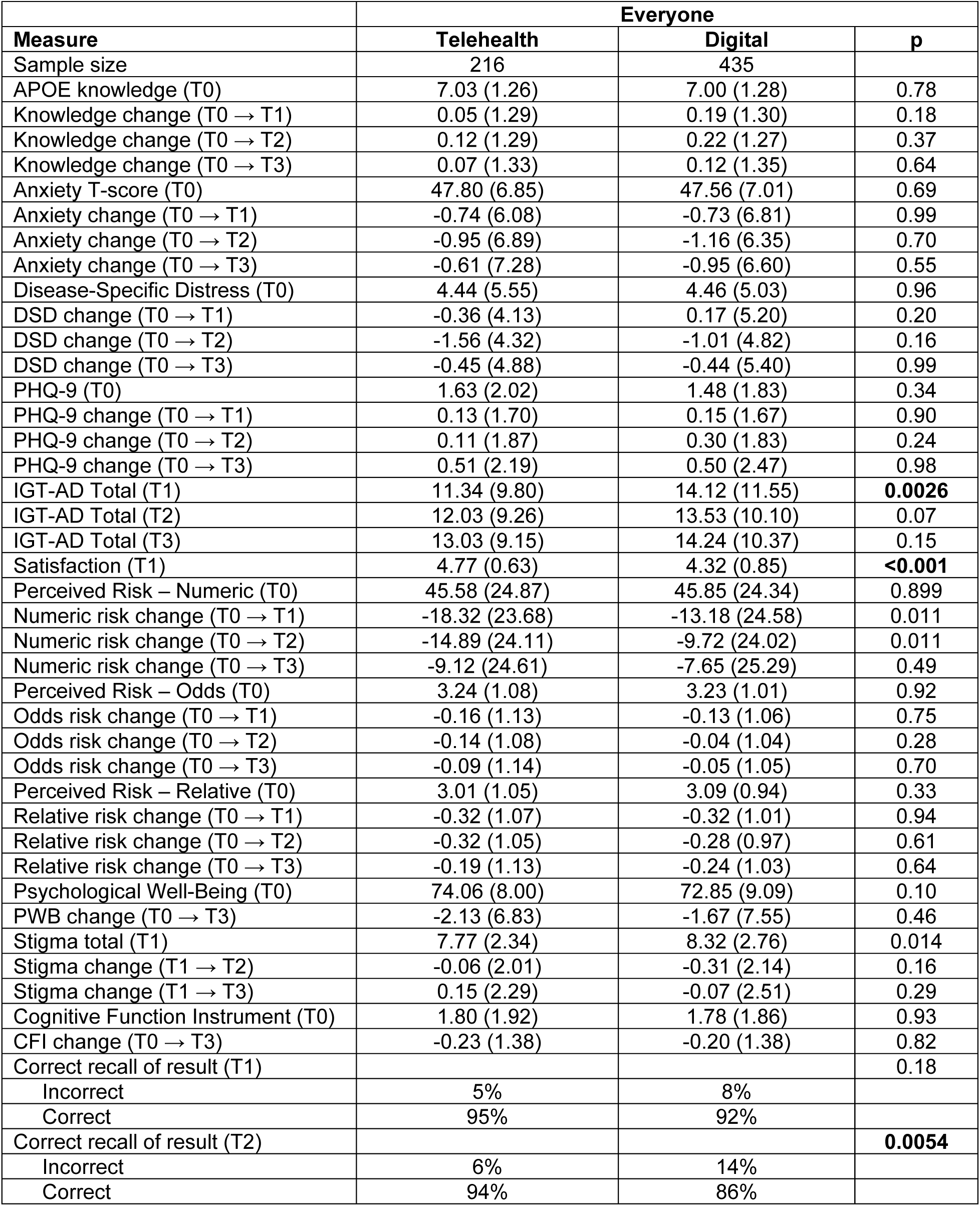
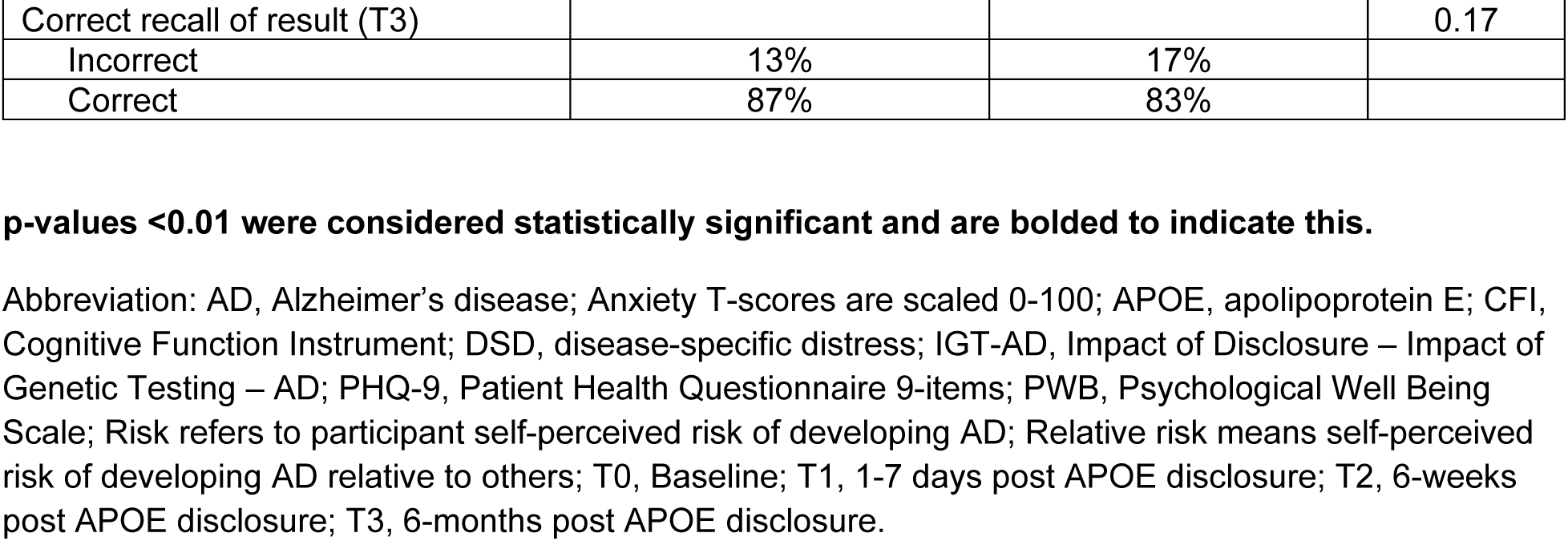
Intention-to-treat analysis by delivery mode for APOE disclosure. Values are presented as mean (Standard Deviation), %, or sample size. This includes the multiply imputed data. p-values <0.01 were considered statistically significant and are bolded to indicate this.

### Outcomes of *APOE* disclosure by arm and by result

Among *APOE4* carriers there were no significant differences between arms for most outcomes. The two exceptions were greater reduction in disease-specific distress in the telehealth arm vs digital arm at T1 (-0.67 [SD 4.3] vs 0.71 [SD 5.4]; *P*=0.0076), although these differences did not persist longitudinally. The overall scores are low and likely subclinical and the differences are small. Thus, these short-term differences are likely not clinically significant. There was also higher negative response to learning *APOE* genetic risk (IGT-AD) at T1 in the digital vs. telehealth arm (17.3 [SD 12.1] vs 14.2 [SD 10.0.008). The differences are small, overall scores are low and are likely not clinically significant. There was also slightly higher satisfaction at T1 in the telehealth vs digital arm (4.7 [SD 0.6] vs 4.3 [SD 0.8]; *P*<0.001), although the differences are small and satisfaction was high in both arms (**Supplemental eTable 3**).

Among non-carriers, there were no significant differences in knowledge or affective outcomes between arms at T1, T2, or T3. Satisfaction was slightly higher in the telehealth vs digital platform arm (4.8 [SD 0.6] vs 4.4 [SD 0.9]; *P*<0.001), although the differences are small and satisfaction was high in both arms (**Supplemental eTable 3**).

### Outcomes of pTau-217 disclosure

As shown in **Table 3**, outcomes did not differ significantly by arm in the ITT analyses. Examining differences by categorical pTau-217 result following disclosure, there was slightly higher satisfaction in the telehealth vs digital arm among those who received an intermediate result (4.6 [SD 0.6] vs 4.3 [SD 0.6]; *P*=0.005), although the differences are small and satisfaction was high in both arms. While there were some cross-sectional differences, the change scores between groups following disclosure were not significant (**Supplemental eTable 4**).

**Table 3.**
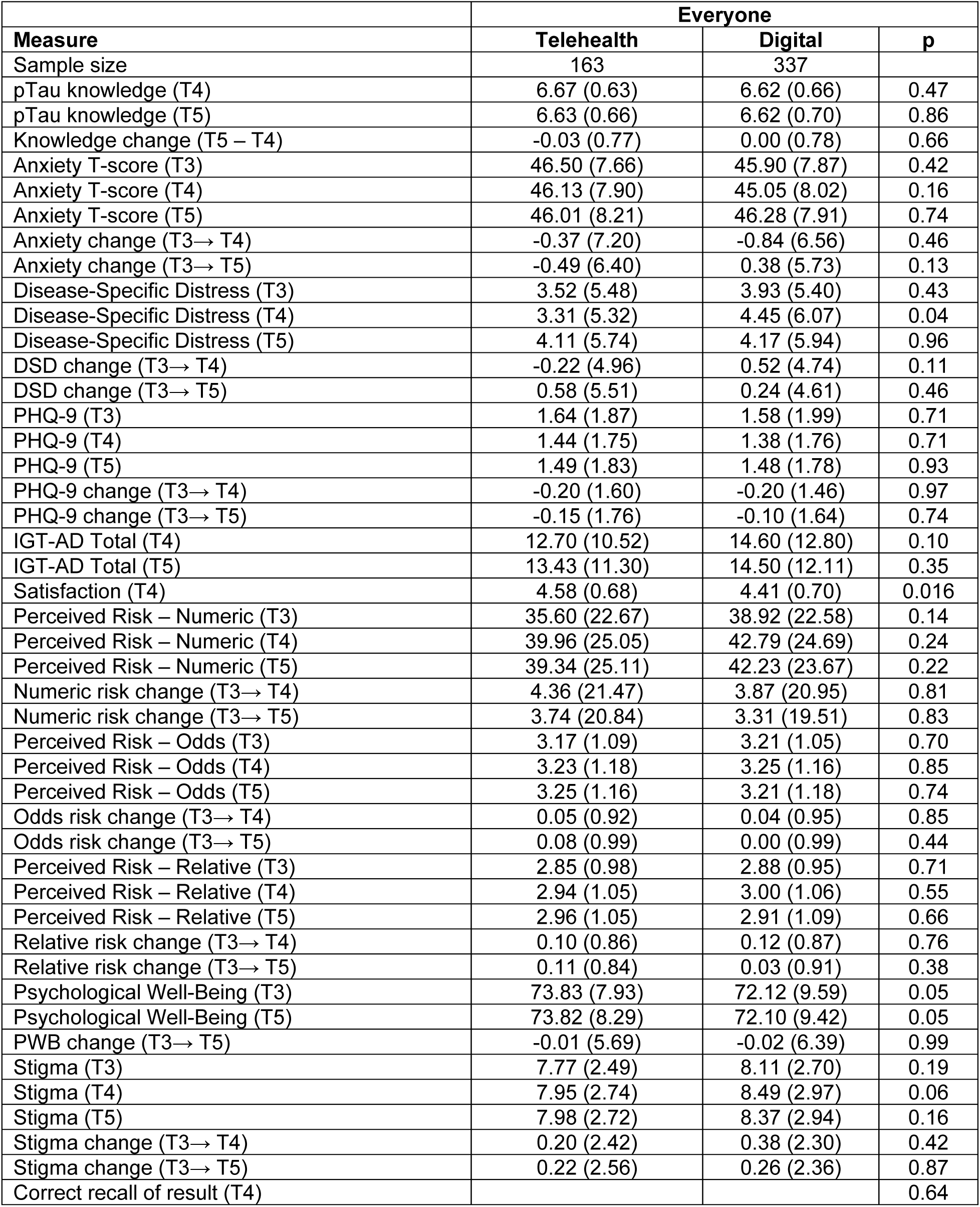
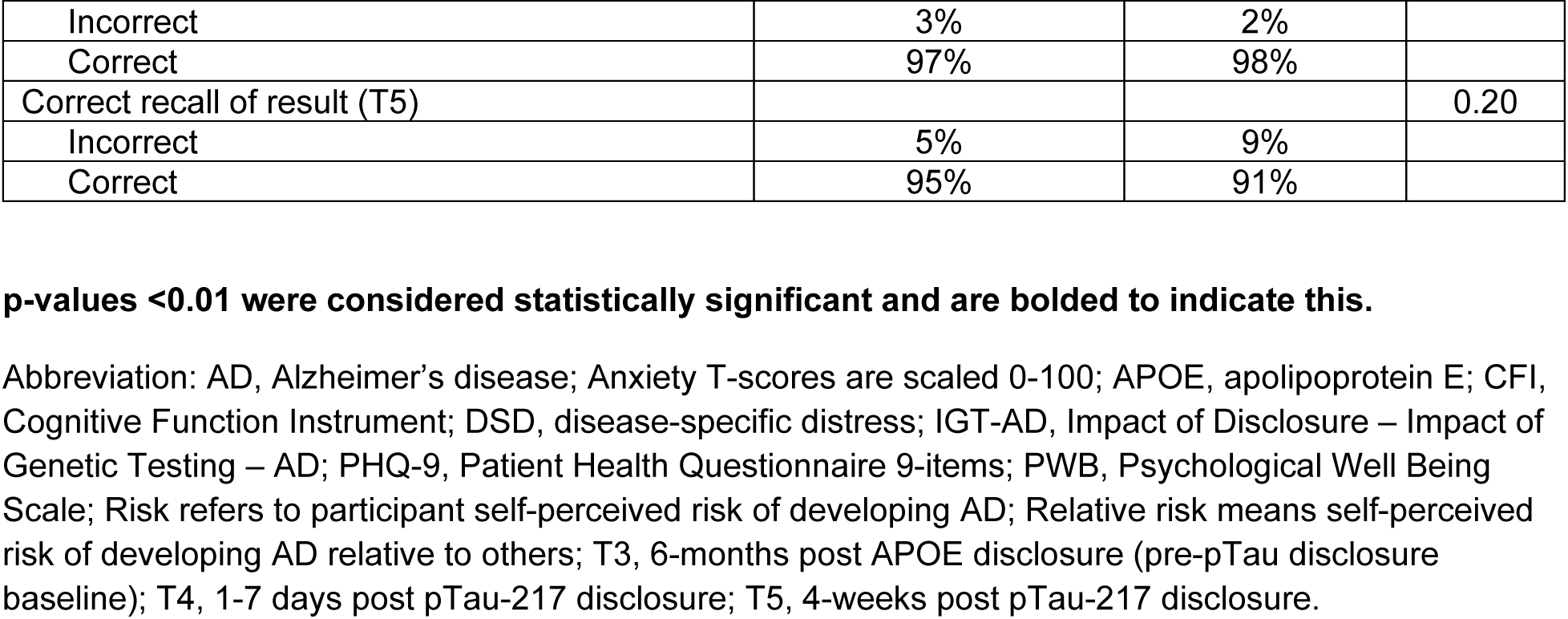
Intention-to-Treat analyses by Delivery Mode for Optional pTau Disclosure: Everyone. Values are presented as mean (Standard Deviation), %, or sample size. This includes the multiply imputed data. p-values <0.01 were considered statistically significant and are bolded to indicate this.

| Measure | Everyone |  |  |
| --- | --- | --- | --- |
|  | Telehealth | Digital | p |
| Sample size | 163 | 337 |  |
| pTau knowledge (T4) | 6.67 (0.63) | 6.62 (0.66) | 0.47 |
| pTau knowledge (T5) | 6.63 (0.66) | 6.62 (0.70) | 0.86 |
| Knowledge change (T5 – T4) | -0.03 (0.77) | 0.00 (0.78) | 0.66 |
| Anxiety T-score (T3) | 46.50 (7.66) | 45.90 (7.87) | 0.42 |
| Anxiety T-score (T4) | 46.13 (7.90) | 45.05 (8.02) | 0.16 |
| Anxiety T-score (T5) | 46.01 (8.21) | 46.28 (7.91) | 0.74 |
| Anxiety change (T3→ T4) | -0.37 (7.20) | -0.84 (6.56) | 0.46 |
| Anxiety change (T3→ T5) | -0.49 (6.40) | 0.38 (5.73) | 0.13 |
| Disease-Specific Distress (T3) | 3.52 (5.48) | 3.93 (5.40) | 0.43 |
| Disease-Specific Distress (T4) | 3.31 (5.32) | 4.45 (6.07) | 0.04 |
| Disease-Specific Distress (T5) | 4.11 (5.74) | 4.17 (5.94) | 0.96 |
| DSD change (T3→ T4) | -0.22 (4.96) | 0.52 (4.74) | 0.11 |
| DSD change (T3→ T5) | 0.58 (5.51) | 0.24 (4.61) | 0.46 |
| PHQ-9 (T3) | 1.64 (1.87) | 1.58 (1.99) | 0.71 |
| PHQ-9 (T4) | 1.44 (1.75) | 1.38 (1.76) | 0.71 |
| PHQ-9 (T5) | 1.49 (1.83) | 1.48 (1.78) | 0.93 |
| PHQ-9 change (T3→ T4) | -0.20 (1.60) | -0.20 (1.46) | 0.97 |
| PHQ-9 change (T3→ T5) | -0.15 (1.76) | -0.10 (1.64) | 0.74 |
| IGT-AD Total (T4) | 12.70 (10.52) | 14.60 (12.80) | 0.10 |
| IGT-AD Total (T5) | 13.43 (11.30) | 14.50 (12.11) | 0.35 |
| Satisfaction (T4) | 4.58 (0.68) | 4.41 (0.70) | 0.016 |
| Perceived Risk – Numeric (T3) | 35.60 (22.67) | 38.92 (22.58) | 0.14 |
| Perceived Risk – Numeric (T4) | 39.96 (25.05) | 42.79 (24.69) | 0.24 |
| Perceived Risk – Numeric (T5) | 39.34 (25.11) | 42.23 (23.67) | 0.22 |
| Numeric risk change (T3→ T4) | 4.36 (21.47) | 3.87 (20.95) | 0.81 |
| Numeric risk change (T3→ T5) | 3.74 (20.84) | 3.31 (19.51) | 0.83 |
| Perceived Risk – Odds (T3) | 3.17 (1.09) | 3.21 (1.05) | 0.70 |
| Perceived Risk – Odds (T4) | 3.23 (1.18) | 3.25 (1.16) | 0.85 |
| Perceived Risk – Odds (T5) | 3.25 (1.16) | 3.21 (1.18) | 0.74 |
| Odds risk change (T3→ T4) | 0.05 (0.92) | 0.04 (0.95) | 0.85 |
| Odds risk change (T3→ T5) | 0.08 (0.99) | 0.00 (0.99) | 0.44 |
| Perceived Risk – Relative (T3) | 2.85 (0.98) | 2.88 (0.95) | 0.71 |
| Perceived Risk – Relative (T4) | 2.94 (1.05) | 3.00 (1.06) | 0.55 |
| Perceived Risk – Relative (T5) | 2.96 (1.05) | 2.91 (1.09) | 0.66 |
| Relative risk change (T3→ T4) | 0.10 (0.86) | 0.12 (0.87) | 0.76 |
| Relative risk change (T3→ T5) | 0.11 (0.84) | 0.03 (0.91) | 0.38 |
| Psychological Well-Being (T3) | 73.83 (7.93) | 72.12 (9.59) | 0.05 |
| Psychological Well-Being (T5) | 73.82 (8.29) | 72.10 (9.42) | 0.05 |
| PWB change (T3→ T5) | -0.01 (5.69) | -0.02 (6.39) | 0.99 |
| Stigma (T3) | 7.77 (2.49) | 8.11 (2.70) | 0.19 |
| Stigma (T4) | 7.95 (2.74) | 8.49 (2.97) | 0.06 |
| Stigma (T5) | 7.98 (2.72) | 8.37 (2.94) | 0.16 |
| Stigma change (T3→ T4) | 0.20 (2.42) | 0.38 (2.30) | 0.42 |
| Stigma change (T3→ T5) | 0.22 (2.56) | 0.26 (2.36) | 0.87 |
| Correct recall of result (T4) |  |  | 0.64 |

|  | Everyone |  |  |
| --- | --- | --- | --- |
| Measure | Telehealth | Digital | p |
| Incorrect | 3% | 2% |  |
| Correct | 97% | 98% |  |
| Correct recall of result (T5) |  |  | 0.20 |
| Incorrect | 5% | 9% |  |
| Correct | 95% | 91% |  |

Abbreviation: AD, Alzheimer's disease; Anxiety T-scores are scaled 0-100; APOE, apolipoprotein E; CFI, Cognitive Function Instrument; DSD, disease-specific distress; IGT-AD, Impact of Disclosure – Impact of Genetic Testing – AD; PHQ-9, Patient Health Questionnaire 9-items; PWB, Psychological Well Being Scale; Risk refers to participant self-perceived risk of developing AD; Relative risk means self-perceived risk of developing AD relative to others; T3, 6-months post APOE disclosure (pre-pTau disclosure baseline); T4, 1-7 days post pTau-217 disclosure; T5, 4-weeks post pTau-217 disclosure.

## Discussion

To our knowledge, this is the first randomized trial to examine whether learning *APOE* results through patient-centered digital platforms is no worse than a telehealth visit with a clinician. It is also the first to explore if outcomes differ between digital and clinician-mediated return of pTau-217 results. Together the results can inform a clinical practice in which patients learn genetic and biomarker results via patient-centered digital platforms, ideally integrated into their electronic health portal, as part of their medical workup for anti-amyloid monoclonal antibody (AAMA) treatments to inform them of their treatment-related risks.

No overall differences in AD-related knowledge, symptoms of anxiety, or disease-specific distress were observed between arms following return of *APOE* results.

However, there were some differences between arms on secondary outcomes and exploratory subgroup analyses. In almost all instances, satisfaction with disclosure services was higher among those in the telehealth arm, although the differences were small. Although there were no differences in recall of *APOE* result between arms immediately (1-7 days) following disclosure nor at 6 months, recall was lower in the digital arm at 6 weeks following disclosure. Higher negative response to genetic testing (IGT-AD) was observed in the digital arm immediately after return of *APOE* results, with no differences at 6 weeks or 6 months after disclosure. Among *APOE4* carriers, disease-specific distress increased slightly and the IGT-AD was higher 1-7 days following disclosure in the digital arm, with no differences observed between arms at 6 weeks and 6 months following disclosure. The observed differences were relatively small, and none of the subgroup mean scores, for measures with clinical thresholds, were in the range that would be considered clinically significant. No differences in knowledge of pTau-217, symptoms of anxiety, disease-specific distress, or secondary outcomes were observed between arms following pTau-217 disclosure. This pattern persisted when stratified by categorical pTau-217 result. The scores observed in the eSMARTER trial align with previous disclosure studies which reported similar changes in mood and wellbeing.^3,5,10,14,17,45^ Together, the results further support that AD-related genetic and biomarker results along with information about a person’s risk of future diagnosis of cognitive impairment due to dementia can be communicated remotely in a safe and effective manner.

We met the a priori defined noninferiority thresholds for outcomes following *APOE* disclosure. In addition, we did not observe differences between arms following disclosure of pTau-217 results. Together, these results suggest that scalable digital platforms are effective methods for communicating these types of sensitive information. This is important as genetic and biomarker testing increase with AAMA therapies and as interest in using biomarker tests in primary care settings expands. Although these platforms are not the same as viewing a lab report in an electronic health portal, there may be opportunities to embed digital platforms into portals as well as for researchers to utilize these platforms in studies without resources to provide clinician-mediated return of results.

## Limitations

Several limitations should be noted. Our study was not designed to assess how return of results via digital platforms compares to learning results from lab reports released to electronic health portals, in which minimal context or education is provided. The digital platforms used in eSMARTER for *APOE* disclosure include information covered by genetic counselors; we recognize that this information is not necessarily discussed by other medical providers or in real-world *APOE* testing. The eSMARTER digital platforms did not offer voice-over narration; participants had to read the text on their screen. This contrasts with the telehealth arm in which clinicians verbally communicated the results. More research is needed to understand the acceptability of digital platforms in returning results for patients with cognitive impairment. Although we tracked whether participants requested to speak with a clinician following their disclosure session or had general questions, the study was not powered to rule out possible differences among subgroups requiring additional or immediate psychological support following disclosure. The sample used is racially homogenous, has a high overall level of education, and self-selected to participate in a decentralized trial, limiting the generalizability of results. Future efforts will explore whether outcomes differ if participants received risk discordant results (e.g., *APOE4* homozygotes with negative pTau-217). The order of results disclosed (*APOE* followed by pTau-217) and the time between disclosure does not reflect current clinical practice.

## Conclusions

In conclusion, this study provides information about the comparability of patient-centered digital platforms and clinician-telehealth sessions for returning AD genetic and biomarker information to adults. The findings support the use of digital platforms for disclosure when speaking with a provider (genetic counselor or clinician) is not available or feasible.^46^ Moreover, these types of digital platforms offer a scalable approach to address the quickly increasing need for returning results to patients at elevated risk for AD dementia.

## Author Contributions

Dr. Langbaum and Dr. Egleston had full access to all the data in the study and take responsibility for the integrity of the data and the accuracy of the data analysis. Data analysis was conducted by Dr. Egleston.

## Additional Contributions

The authors would like to extend their gratitude the site principal investigators, staff, and participants for their involvement in the study. We also thank Dr. Joshua Grill for serving as the study’s safety officer, as well as Dr. Robert Alexlander, Dr. Tainá M. Marques, Kari Dieckhoff, Davron Hanley, Darlene Ellenor, and James Liu for their assistance during the conduct of the study.

## Data Availability

Deidentified data are available. Requests for access to the study data can be submitted by emailing a description of your request to. A Data Use Agreement will need to be completed with Banner Health prior to transfer of data.

