## Supplemental Materials for "Returning *APOE* and pTau-217 Results: the eSMARTER Randomized Noninferiority Clinical Trial"

### 1. Table of Contents: Supplementary Materials

**eTable 1: eSMARTER assessments**

| eSMARTER Assessment | Brief Description | Score Range | Timepoints Assessed |
|---|---|---|---|
| Demographics and Clinical Characterization Questions | Includes date of birth, sex, gender, address, race, ethnicity, years of education, employment status, income, sexual orientation, marital status, living situation, family history of AD, and history of mild cognitive impairment (MCI) or dementia diagnosis. | Not Applicable | Screening, T0 |
| Abbreviated Columbia Suicide Severity Scale (C-SSRS) & AD Specific Suicidality Questions | 4-item self-report assessment of general and AD-specific suicidality using 2 questions from the C-SSRS and 2 questions from the Alzheimer's Disease Neuroimaging Initiative (ADNI) 4 study specific to learning high risk of AD. | 0 – 4 | Screening, T3 |
| Patient Health Questionnaire – 9 (PHQ-9) | 9-item self-report assessment of depressive symptoms over the past 2 weeks. A score of 10 or greater, indicating at least moderate depression, was exclusionary. | 0 – 27 | Screening, T0, T1, T2, T3, T4, T5 |
| Prior Knowledge of APOE Status | Single self-report item assessing whether a participant already knows their APOE genetic status. | Not Applicable | Screening |
| Telephone Montreal Cognitive Assessment (T-MoCA) | Modified 15-item version of the MoCA for use over the phone to assess general cognitive status. Administered by eSMARTER study coordinators over the phone. | 0 - 22 | Screening |
| Health Literacy | 3-item self-report assessment of how well the participant understands written information relating to their health. | 0 - 12 | T0 |
| Comfort with Technology | Select self-report questions extracted from the Health Information National Trends Survey (HINTS) to assess comfort with health-related technology. | Not Applicable | T0 |
| Knowledge of APOE | 8-item self-report scale adapted from the Cancer Genetics Knowledge Scale and ClinSeq Knowledge Scale to to evaluate genetic knowledge in the context of APOE testing for AD. | 0 - 8 | T0, T1, T2, T3 |
| Disclosure Modality Preference | Single self-report item rank ordering preference of modality to receive AD related risk information and results. | Not Applicable | T0, T1, T4 |
| Satisfaction with Disclosure Services | 15-item self-report scale evaluating the participants' comfort and satisfaction with the length and modality they used to obtain the genetic information and feelings/emotions resulting from the session | 0 - 70 | T1, T4 |
| System Usability Scale (SUS) | Modified version of the self-reported 10-item System Usability Scale for use with cognitively impaired older adults to assess usability of the eSMARTER platforms. | 0 - 100 | T1, T4 |

| | | | |
|---|---|---|---|
| Psychological Wellbeing Scale – Modified Version | 14 self-report items from the abbreviated 18-item version of this scale, which measures six aspects of wellbeing and happiness. | 0 - 84 | T0, T3, T5 |
| Disease Specific Distress | Eight self-report items adapted from the Impact of Events Scale (IES) to evaluate disease-specific distress among both adolescents and adults. | 0 - 40 | T0, T1, T2, T3, T4, T5 |
| PROMIS Anxiety | 7-item self-report anxiety short-form version PROMIS (Patient-Reported Outcomes Measurement Information System) to assess anxious symptoms. | 7 - 35 | T0, T1, T2, T3, T4, T5 |
| Perceived Risk of AD | 3-item self-report assessment of individuals' beliefs regarding their likelihood of developing Alzheimer's disease in the future. | Not Applicable | T0, T1, T2, T3, T4, T5 |
| Impact of Disclosure – Impact of Genetic Testing – AD (IGT-AD) | 16-item self-report measure consisting of two subscales: a) 12-item distress scale that assesses psychological distress in response to learning one's genetic test results, and b) a 4-item positive response subscale that assesses psychological benefits (e.g., relief from anxiety) of learning one's genetic testing results. | 0 - 80 | T1, T2, T3, T4, T5 |
| Recall of Result – APOE | Single self-report item assessing accurate recall of the participant's APOE genetic test result provided at their disclosure session. | 0 - 2 | T1, T2, T3 |
| Cognitive Function Instrument (CFI) | 14-item self-reported outcome to evaluate cognitive and functional decline. | 0 - 14 | T0, T3 |
| Sharing Result with Others | Self-report questionnaire developed by the study team asking participants to report with whom they have shared their AD risk results. | Not Applicable | T3, T5 |
| Modified Stigma Impact Scale (mSIS) | 6 self-report questions from the Stigma Impact Scale measuring impact of negative social attitudes toward the patients' health or mental health condition or disorder | 6 - 24 | T1, T2, T3, T4, T5 |
| Health Behavior Changes | A self-report questionnaire developed by the study team asking participants to report on whether they are making or considering changes to a variety of areas impacting health. | Not Applicable | T3, T5 |
| Knowledge of pTau-217 | A 5-item assessment developed by the study team asking participants to answer "True", "False", or "Not Sure" to questions designed to assess the participants knowledge level of pTau-217. | 0 - 5 | T4, T5 |
| Recall of pTau-217 result | Single self-report item assessing accurate recall of the participant's pTau-217 test result provided at their disclosure session. | 0 - 1 | |

Abbreviation: AD, Alzheimer's disease; APOE, apolipoprotein E; T0, Baseline; T1, 1-7 days post APOE disclosure; T2, 6-weeks post APOE disclosure; T3, 6-months post APOE disclosure; T4, 1-7 days post pTau-217 disclosure; T5, 4-weeks post pTau-217 disclosure

**eTable 2. Sample characteristics by retention post-screening**

Missing values are reported but were not used to calculate p-values.

| | APOE Disclosed (N = 651) | Withdrew or lost to follow-up after screening (N = 23) | Total (N = 674) | p-value |
|---|---|---|---|---|
| **Age** | 67.90 (4.70) | 69.87 (4.72) | 67.96 (4.71) | 0.048 |
| **Sex** | | | | 0.013 |
| Female | 439 (67.4%) | 12 (52.2%) | 451 (66.9%) | |
| Male | | | | |
| **Race** | | | | 0.09 |
| American Indian or Alaska Native | 4 (0.6%) | 1 (4.8%) | 5 (0.8%) | |
| Asian | 13 (2.0%) | 0 (0.0%) | 13 (2.0%) | |
| Black or African American | 21 (3.3%) | 2 (9.5%) | 23 (3.5%) | |
| White | 585 (91.1%) | 18 (85.7%) | 603 (91.0%) | |
| Other | 19 (3.0%) | 0 (0.0%) | 19 (2.9%) | |
| Missing | 9 (.%) | 2 (.%) | 11 (.%) | |
| **Ethnicity** | | | | 0.024 |
| Not Hispanic | 613 (94.2%) | 19 (82.6%) | 632 (93.8%) | |
| Hispanic | 38 (5.8%) | 4 (17.4%) | 42 (6.2%) | |
| **Marital Status** | | | | 0.55 |
| Currently married or partnered | 468 (72.0%) | 14 (87.5%) | 482 (72.4%) | |
| Widowed | 57 (8.8%) | 1 (6.2%) | 58 (8.7%) | |
| Divorced or Separated | 95 (14.6%) | 1 (6.2%) | 96 (14.4%) | |
| Never Married | 30 (4.6%) | 0 (0.0%) | 30 (4.5%) | |
| Missing | 1 (.%) | 7 (.%) | 8 (.%) | |
| **Educational Level** | | | | 0.032 |
| High School or less | 24 (3.7%) | 0 (0.0%) | 24 (3.6%) | |
| Some College or Trade School | 119 (18.3%) | 7 (43.8%) | 126 (18.9%) | |
| College or more | 507 (78.0%) | 9 (56.2%) | 516 (77.5%) | |
| Missing | 1 (.%) | 7 (.%) | 8 (.%) | |
| **Family History of Alzheimer's** | | | | 0.74 |
| No | 129 (19.8%) | 2 (12.5%) | 131 (19.7%) | |
| Yes | 434 (66.8%) | 12 (75.0%) | 446 (67.0%) | |
| Not sure | 87 (13.4%) | 2 (12.5%) | 89 (13.4%) | |
| Missing | 1 (.%) | 7 (.%) | 8 (.%) | |
| **Telephone MoCA Total Score (0-22)** | 19.00 (2.00) | 18.30 (1.94) | 18.98 (2.00) | 0.10 |
| **ADI National Rank (1-100)** | 38.54 (23.80) | 41.29 (15.91) | 38.61 (23.63) | 0.64 |
| **Rurality** | | | | 0.06 |
| City/Suburban/Town | 308 (47.8%) | 12 (70.6%) | 320 (48.4%) | |
| Rural | 336 (52.2%) | 5 (29.4%) | 341 (51.6%) | |
| Missing | 7 (.%) | 6 (.%) | 13 (.%) | |
| **Income** | | | | 0.07 |
| <$100,000 | 329 (54.3%) | 11 (78.6%) | 340 (54.8%) | |

| | APOE Disclosed (N = 651) | Withdrew or lost to follow-up after screening (N = 23) | Total (N = 674) | p-value |
|---|---|---|---|---|
| >=$100,000 | 277 (45.7%) | 3 (21.4%) | 280 (45.2%) | |
| Missing | 45 (.%) | 9 (.%) | 54 (.%) | |
| **Employment Status** | | | | 0.70 |
| Employed or Homemaker | 207 (31.8%) | 6 (37.5%) | 213 (32.0%) | |
| Not Working | 22 (3.4%) | 0 (0.0%) | 22 (3.3%) | |
| Retired | 421 (64.8%) | 10 (62.5%) | 431 (64.7%) | |
| Missing | 1 (.%) | 7 (.%) | 8 (.%) | |
| **APOE Carrier Status** | | | | 0.12 |
| Homozygote | 66 (10.1%) | 2 (8.7%) | 68 (10.1%) | |
| Heterozygote | 377 (57.9%) | 9 (39.1%) | 386 (57.3%) | |
| Non-Carrier | 208 (32.0%) | 12 (52.2%) | 220 (32.6%) | |

Abbreviation: ADI, Area Deprivation Index; APOE, apolipoprotein E; MoCA, Montreal Cognitive Assessment

**eTable 3. Intention-to-treat analysis by delivery mode across APOE groups**

Values are presented as mean (Standard Deviation), %, or sample size. This includes the multiply imputed data. P-values compare arms within each APOE subgroup.

| | **APOE e4 Carriers (homozygotes and heterozygotes)** | | | **APOE e4 Non-Carriers** | | |
|---|---|---|---|---|---|---|
| **Measure** | **Telehealth** | **Digital** | **p** | **Telehealth** | **Digital** | **p** |
| Sample size | 146 | 297 | | 70 | 138 | |
| APOE knowledge (T0) | 7.08 (1.20) | 7.03 (1.20) | 0.70 | 6.94 (1.38) | 6.94 (1.45) | 0.99 |
| Knowledge change (T0 → T1) | -0.02 (1.24) | 0.15 (1.26) | 0.18 | 0.19 (1.39) | 0.28 (1.39) | 0.65 |
| Knowledge change (T0 → T2) | 0.09 (1.17) | 0.19 (1.23) | 0.40 | 0.20 (1.52) | 0.28 (1.36) | 0.69 |
| Knowledge change (T0 → T3) | 0.06 (1.22) | 0.06 (1.30) | 0.96 | 0.09 (1.53) | 0.25 (1.44) | 0.48 |
| Anxiety T-score (T0) | 47.71 (6.27) | 47.71 (6.88) | 1.00 | 47.98 (7.98) | 47.25 (7.31) | 0.51 |
| Anxiety change (T0 → T1) | -1.21 (6.04) | -0.45 (7.05) | 0.27 | 0.23 (6.10) | -1.34 (6.24) | 0.09 |
| Anxiety change (T0 → T2) | -1.59 (6.74) | -1.20 (6.51) | 0.56 | 0.39 (7.05) | -1.08 (6.02) | 0.13 |
| Anxiety change (T0 → T3) | -1.17 (6.71) | -1.32 (6.43) | 0.82 | 0.57 (8.26) | -0.14 (6.90) | 0.52 |
| Disease-Specific Distress (T0) | 4.93 (5.59) | 4.84 (5.19) | 0.86 | 3.40 (5.35) | 3.63 (4.57) | 0.75 |
| DSD change (T0 → T1) | -0.67 (4.27) | 0.71 (5.36) | **0.0076** | 0.31 (3.77) | -0.99 (4.63) | 0.046 |
| DSD change (T0 → T2) | -1.73 (4.36) | -0.79 (5.02) | 0.06 | -1.21 (4.26) | -1.47 (4.33) | 0.69 |
| DSD change (T0 → T3) | -0.55 (5.19) | -0.48 (5.31) | 0.89 | -0.22 (4.16) | -0.35 (5.61) | 0.87 |
| PHQ-9 (T0) | 1.62 (2.00) | 1.39 (1.78) | 0.22 | 1.66 (2.06) | 1.68 (1.92) | 0.93 |
| PHQ-9 change (T0 → T1) | -0.05 (1.68) | 0.12 (1.54) | 0.30 | 0.50 (1.68) | 0.21 (1.92) | 0.29 |
| PHQ-9 change (T0 → T2) | 0.16 (1.88) | 0.30 (1.79) | 0.46 | 0.00 (1.84) | 0.29 (1.94) | 0.32 |
| PHQ-9 change (T0 → T3) | 0.37 (2.24) | 0.34 (2.14) | 0.89 | 0.81 (2.06) | 0.87 (3.05) | 0.89 |
| IGT-AD Total (T1) | 14.19 (9.95) | 17.29 (12.13) | **0.008** | 5.38 (6.17) | 7.28 (6.01) | 0.039 |
| IGT-AD Total (T2) | 14.65 (9.37) | 15.97 (10.66) | 0.21 | 6.56 (6.14) | 8.30 (6.09) | 0.06 |
| IGT-AD Total (T3) | 15.25 (9.31) | 16.58 (10.79) | 0.21 | 8.39 (6.81) | 9.22 (7.20) | 0.44 |
| Satisfaction (T1) | 4.74 (0.61) | 4.30 (0.81) | **<0.001** | 4.84 (0.66) | 4.35 (0.91) | **<0.001** |
| Perceived Risk – Numeric (T0) | 48.48 (24.95) | 48.26 (23.36) | 0.93 | 39.54 (23.76) | 40.66 (25.66) | 0.76 |
| Numeric risk change (T0 → T1) | -17.39 (24.38) | -11.81 (24.43) | 0.025 | -20.28 (22.20) | -16.13 (24.70) | 0.24 |
| Numeric risk change (T0 → T2) | -14.02 (25.06) | -8.86 (23.30) | 0.037 | -16.70 (22.03) | -11.58 (25.48) | 0.16 |
| Numeric risk change (T0 → T3) | -7.76 (25.94) | -6.29 (24.53) | 0.57 | -11.95 (21.47) | -10.58 (26.72) | 0.72 |
| Perceived Risk – Odds (T0) | 3.34 (1.10) | 3.35 (0.94) | 0.97 | 3.03 (1.01) | 2.99 (1.10) | 0.78 |

| | APOE e4 Carriers (homozygotes and heterozygotes) | | | APOE e4 Non-Carriers | | |
|---|---|---|---|---|---|---|
| **Measure** | **Telehealth** | **Digital** | **p** | **Telehealth** | **Digital** | **p** |
| Odds risk change (T0 → T1) | 0.11 (1.10) | 0.09 (1.02) | 0.88 | -0.70 (0.98) | -0.59 (1.02) | 0.46 |
| Odds risk change (T0 → T2) | 0.12 (1.03) | 0.13 (1.03) | 0.92 | -0.67 (1.00) | -0.39 (0.97) | 0.07 |
| Odds risk change (T0 → T3) | 0.11 (1.11) | 0.12 (1.02) | 0.93 | -0.50 (1.10) | -0.42 (1.03) | 0.62 |
| Perceived Risk – Relative (T0) | 3.14 (1.04) | 3.22 (0.89) | 0.43 | 2.74 (1.02) | 2.83 (1.00) | 0.57 |
| Relative risk change (T0 → T1) | -0.19 (1.08) | -0.19 (1.02) | 0.98 | -0.58 (1.01) | -0.60 (0.94) | 0.87 |
| Relative risk change (T0 → T2) | -0.18 (1.02) | -0.17 (0.95) | 0.90 | -0.62 (1.05) | -0.52 (0.97) | 0.51 |
| Relative risk change (T0 → T3) | -0.08 (1.14) | -0.13 (1.01) | 0.67 | -0.41 (1.08) | -0.46 (1.04) | 0.78 |
| Psychological Well-Being (T0) | 73.74 (8.08) | 73.02 (8.31) | 0.39 | 74.74 (7.83) | 72.48 (10.60) | 0.12 |
| PWB change (T0 → T3) | -1.66 (6.99) | -1.37 (7.46) | 0.71 | -3.12 (6.40) | -2.31 (7.73) | 0.48 |
| Stigma total (T1) | 8.26 (2.53) | 8.83 (2.94) | 0.049 | 6.76 (1.45) | 7.23 (1.92) | 0.08 |
| Stigma change (T1→ T2) | -0.06 (2.27) | -0.34 (2.25) | 0.24 | -0.07 (1.33) | -0.26 (1.86) | 0.46 |
| Stigma change (T1→ T3) | 0.14 (2.54) | -0.17 (2.55) | 0.25 | 0.17 (1.65) | 0.13 (2.42) | 0.91 |
| Cognitive Function Instrument (T0) | 1.79 (1.90) | 1.80 (1.84) | 0.99 | 1.80 (1.97) | 1.75 (1.89) | 0.86 |
| CFI change (T0 → T3) | -0.27 (1.38) | -0.23 (1.37) | 0.79 | -0.14 (1.38) | -0.14 (1.38) | 0.98 |
| Correct recall of result (T1) | | | 0.30 | | | 0.40 |
| Incorrect | 5% | 7% | | 5% | 9% | |
| Correct | 95% | 93% | | 95% | 91% | |
| Correct recall of result (T2) | | | 0.025 | | | 0.13 |
| Incorrect | 7% | 15% | | 3% | 11% | |
| Correct | 93% | 85% | | 97% | 89% | |
| Correct recall of result (T3) | | | 0.29 | | | 0.37 |
| Incorrect | 14% | 18% | | 12% | 17% | |
| Correct | 86% | 82% | | 88% | 83% | |

p-values <0.01 were considered statistically significant and are bolded to indicate this.

Abbreviation: AD, Alzheimer's disease; Anxiety T-scores are scaled 0-100; APOE, apolipoprotein E; CFI, Cognitive Function Instrument; DSD, disease-specific distress; IGT-AD, Impact of Disclosure – Impact of Genetic Testing – AD; PHQ-9, Patient Health Questionnaire 9-items; PWB, Psychological Well Being Scale; Risk refers to participant self-perceived risk of developing AD; Relative risk means self-perceived risk of developing AD relative to others; T0, Baseline; T1, 1-7 days post APOE disclosure; T2, 6-weeks post APOE disclosure; T3, 6-months post APOE disclosure.

**eTable 4. Intention-to-treat analysis by delivery mode across pTau-217 subgroups**

Values are presented as mean (Standard Deviation), %, or sample size. This includes the multiply imputed data. P-values compare arms within each pTau-217 subgroup.

| | pTau-217 Positive | | | pTau-217 Intermediate | | | pTau-217 Negative | | |
|---|---|---|---|---|---|---|---|---|---|
| **Measure** | **Telehealth** | **Digital** | **p** | **Telehealth** | **Digital** | **p** | **Telehealth** | **Digital** | **p** |
| Sample size | 43 | 67 | | 45 | 108 | | 75 | 162 | |
| pTau knowledge (T4) | 6.63 (0.62) | 6.69 (0.63) | 0.61 | 6.73 (0.58) | 6.63 (0.63) | 0.37 | 6.65 (0.67) | 6.59 (0.69) | 0.50 |
| pTau knowledge (T5) | 6.71 (0.55) | 6.45 (0.95) | 0.11 | 6.67 (0.56) | 6.69 (0.65) | 0.86 | 6.57 (0.76) | 6.65 (0.59) | 0.38 |
| Knowledge change (T0→ T5) | 0.08 (0.64) | -0.24 (0.90) | 0.05 | -0.07 (0.83) | 0.05 (0.68) | 0.36 | -0.08 (0.80) | 0.06 (0.78) | 0.20 |
| Anxiety T-score (T0) | 47.14 (7.28) | 46.45 (8.40) | 0.66 | 46.27 (7.53) | 45.31 (7.18) | 0.46 | 46.27 (8.03) | 46.06 (8.11) | 0.85 |
| Anxiety T-score (T4) | 47.96 (8.01) | 48.17 (8.65) | 0.90 | 44.67 (7.71) | 44.49 (7.93) | 0.90 | 45.97 (7.84) | 44.14 (7.54) | 0.09 |
| Anxiety T-score (T5) | 48.23 (9.03) | 47.87 (8.05) | 0.83 | 44.76 (8.28) | 45.71 (7.76) | 0.50 | 45.48 (7.51) | 45.99 (7.92) | 0.64 |
| Anxiety change (T0→ T4) | 0.82 (7.21) | 1.72 (7.10) | 0.52 | -1.61 (6.27) | -0.82 (6.59) | 0.50 | -0.30 (7.68) | -1.92 (6.03) | 0.08 |
| Anxiety change (T0→ T5) | 1.09 (6.85) | 1.43 (6.44) | 0.80 | -1.52 (5.84) | 0.40 (5.67) | 0.07 | -0.79 (6.36) | -0.07 (5.43) | 0.38 |
| Disease-Specific Distress (T0) | 3.83 (5.82) | 5.42 (6.55) | 0.20 | 3.40 (6.35) | 3.87 (4.98) | 0.63 | 3.43 (4.73) | 3.36 (5.05) | 0.93 |
| Disease-Specific Distress (T4) | 4.33 (5.81) | 8.15 (8.31) | **0.010** | 3.02 (5.60) | 4.44 (5.54) | 0.15 | 2.89 (4.82) | 2.93 (4.50) | 0.95 |
| Disease-Specific Distress (T5) | 6.09 (6.25) | 8.05 (8.64) | 0.20 | 4.71 (7.06) | 3.84 (5.00) | 0.39 | 2.61 (3.94) | 2.78 (4.27) | 0.77 |
| DSD change (T0→ T4) | 0.50 (6.50) | 2.73 (5.63) | 0.06 | -0.38 (5.05) | 0.57 (4.82) | 0.28 | -0.53 (3.77) | -0.43 (3.94) | 0.85 |
| DSD change (T0→ T5) | 2.26 (6.83) | 2.63 (5.77) | 0.76 | 1.31 (6.04) | -0.03(4.82) | 0.16 | -0.81 (3.78) | -0.58 (3.49) | 0.64 |
| PHQ-9 (T0) | 1.72 (2.17) | 1.58 (1.98) | 0.73 | 1.47 (1.36) | 1.43 (1.84) | 0.89 | 1.71 (1.96) | 1.67 (2.10) | 0.91 |
| PHQ-9 (T4) | 1.58 (1.89) | 1.34 (1.56) | 0.49 | 1.62 (1.89) | 1.34 (1.81) | 0.39 | 1.25 (1.57) | 1.41 (1.82) | 0.51 |
| PHQ-9 (T5) | 2.02 (2.34) | 1.47 (1.80) | 0.17 | 1.47 (1.58) | 1.51 (1.82) | 0.89 | 1.20 (1.56) | 1.46 (1.75) | 0.28 |

| | pTau-217 Positive | | | pTau-217 Intermediate | | | pTau-217 Negative | | |
|---|---|---|---|---|---|---|---|---|---|
| **Measure** | **Telehealth** | **Digital** | **p** | **Telehealth** | **Digital** | **p** | **Telehealth** | **Digital** | **p** |
| Sample size | 43 | 67 | | 45 | 108 | | 75 | 162 | |
| PHQ-9 change (T0→ T4) | -0.14 (1.40) | -0.24 (1.54) | 0.75 | 0.16 (1.43) | -0.08 (1.42) | 0.35 | -0.45 (1.77) | -0.26 (1.46) | 0.37 |
| PHQ-9 change (T0→ T5) | 0.30 (2.09) | -0.12 (1.59) | 0.25 | 0.00 (1.16) | 0.09 (1.53) | 0.75 | -0.51 (1.80) | -0.22 (1.72) | 0.24 |
| IGT-AD Total (T4) | 21.12 (11.29) | 26.04 (14.17) | 0.06 | 14.29 (9.44) | 17.51 (11.99) | 0.11 | 6.92 (6.33) | 7.93 (7.68) | 0.32 |
| IGT-AD Total (T5) | 21.67 (12.29) | 24.84 (14.68) | 0.24 | 15.82 (11.38) | 16.53 (10.40) | 0.71 | 7.27 (5.98) | 8.87 (8.17) | 0.14 |
| Satisfaction (T4) | 4.49 (0.67) | 4.41 (0.69) | 0.58 | 4.64 (0.61) | 4.32 (0.65) | **0.005** | 4.59 (0.72) | 4.48 (0.73) | 0.29 |
| Perceived Risk – Numeric (T0) | 35.36 (21.17) | 40.30 (21.06) | 0.24 | 37.73 (22.72) | 40.40 (20.54) | 0.48 | 34.45 (23.65) | 37.36 (24.43) | 0.39 |
| Perceived Risk – Numeric (T4) | 53.20 (25.49) | 60.90 (22.15) | 0.10 | 42.42 (24.51) | 46.47 (22.18) | 0.32 | 30.89 (21.42) | 32.85 (22.33) | 0.53 |
| Perceived Risk – Numeric (T5) | 50.25 (27.45) | 57.13 (23.72) | 0.17 | 41.36 (25.02) | 46.07 (21.06) | 0.25 | 31.86 (21.24) | 33.50 (21.55) | 0.59 |
| Numeric risk change (T0→ T4) | 17.84 (24.85) | 20.60 (21.98) | 0.55 | 4.69 (18.71) | 6.08 (19.39) | 0.69 | -3.56 (16.77) | -4.51 (16.64) | 0.69 |
| Numeric risk change (T0→ T5) | 14.89 (28.45) | 16.83 (22.23) | 0.70 | 3.63 (15.45) | 5.68 (16.74) | 0.50 | -2.59 (15.37) | -3.85 (16.56) | 0.59 |
| Perceived Risk – Odds (T0) | 3.16 (1.06) | 3.34 (0.91) | 0.35 | 3.29 (1.04) | 3.35 (0.99) | 0.75 | 3.11 (1.13) | 3.07 (1.11) | 0.80 |
| Perceived Risk – Odds (T4) | 3.81 (1.02) | 4.07 (0.86) | 0.16 | 3.44 (1.01) | 3.48 (1.04) | 0.86 | 2.76 (1.18) | 2.75 (1.10) | 0.95 |
| Perceived Risk – Odds (T5) | 3.84 (0.99) | 3.91 (0.97) | 0.72 | 3.37 (1.08) | 3.39 (1.07) | 0.92 | 2.84 (1.14) | 2.80 (1.17) | 0.81 |
| Odds risk change (T0→ T4) | 0.64 (1.02) | 0.73 (0.94) | 0.68 | 0.16 (0.67) | 0.13 (0.91) | 0.89 | -0.35 (0.80) | -0.32 (0.80) | 0.78 |
| Odds risk change (T0→ T5) | 0.68 (1.00) | 0.57 (1.12) | 0.64 | 0.08 (0.85) | 0.04 (0.93) | 0.84 | -0.27 (0.90) | -0.27 (0.86) | 1.00 |
| Perceived Risk – Relative (T0) | 2.84 (0.94) | 3.00 (0.94) | 0.39 | 3.02 (0.96) | 2.97 (0.89) | 0.74 | 2.75 (1.01) | 2.77 (0.98) | 0.85 |
| Perceived Risk – Relative (T4) | 3.50 (1.00) | 3.76 (0.84) | 0.18 | 3.20 (0.84) | 3.20 (0.93) | 0.98 | 2.47 (0.98) | 2.56 (1.00) | 0.52 |
| Perceived Risk – Relative (T5) | 3.55 (0.88) | 3.55 (0.96) | 0.96 | 3.04 (1.06) | 3.03 (1.00) | 0.99 | 2.56 (0.97) | 2.56 (1.06) | 0.98 |
| Relative risk change | 0.66 (0.93) | 0.76 (0.92) | 0.62 | 0.18 (0.58) | 0.24 (0.85) | 0.68 | -0.28 | -0.22 | 0.54 |

| | pTau-217 Positive | | | pTau-217 Intermediate | | | pTau-217 Negative | | |
|---|---|---|---|---|---|---|---|---|---|
| **Measure** | **Telehealth** | **Digital** | **p** | **Telehealth** | **Digital** | **p** | **Telehealth** | **Digital** | **p** |
| Sample size | 43 | 67 | | 45 | 108 | | 75 | 162 | |
| (T0→ T4) | | | | | | | (0.78) | (0.68) | |
| Relative risk change (T0→ T5) | 0.72 (0.89) | 0.55 (1.01) | 0.40 | 0.01 (0.68) | 0.07 (0.96) | 0.77 | -0.18 (0.72) | -0.21 (0.71) | 0.78 |
| Psychological Well-Being (T0) | 73.58 (7.66) | 74.11 (7.51) | 0.73 | 73.70 (8.61) | 72.66 (10.31) | 0.55 | 74.04 (7.75) | 70.94 (9.74) | 0.017 |
| Psychological Well-Being (T5) | 72.99 (7.86) | 73.73 (7.06) | 0.61 | 73.60 (9.69) | 72.85 (9.94) | 0.68 | 74.42 (7.67) | 70.92 (9.80) | **0.008** |
| PWB change (T0→ T5) | -0.59 (5.43) | -0.37 (6.18) | 0.86 | -0.10 (6.63) | 0.20 (6.96) | 0.82 | 0.38 (5.27) | -0.02 (6.11) | 0.64 |
| Stigma (T0) | 7.65 (2.30) | 9.22 (3.10) | **0.005** | 8.25 (2.82) | 7.90 (2.29) | 0.42 | 7.56 (2.37) | 7.78 (2.67) | 0.55 |
| Stigma (T4) | 9.18 (3.38) | 10.30 (3.45) | 0.10 | 8.53 (3.00) | 8.94 (2.86) | 0.43 | 6.91 (1.56) | 7.43 (2.33) | 0.08 |
| Stigma (T5) | 9.12 (3.51) | 10.11 (3.54) | 0.16 | 8.55 (2.69) | 8.88 (2.85) | 0.51 | 6.99 (1.71) | 7.31 (2.21) | 0.28 |
| Stigma change (T0→ T4) | 1.53 (2.56) | 1.07 (2.44) | 0.36 | 0.29 (2.27) | 1.05 (2.45) | 0.08 | -0.62 (2.09) | -0.36 (1.89) | 0.34 |
| Stigma change (T0→ T5) | 1.46 (2.99) | 0.88 (2.48) | 0.28 | 0.31 (2.32) | 0.99 (2.40) | 0.11 | -0.54 (2.15) | -0.48 (2.06) | 0.84 |
| Correct recall of result (T4) | | | 0.19 | | | 1.00 | | | 0.81 |
| Incorrect | 10% | 3% | | 0% | 3% | | 1% | 2% | |
| Correct | 90% | 97% | | 100% | 97% | | 99% | 98% | |
| Correct recall of result (T5) | | | 0.72 | | | 0.60 | | | 0.25 |
| Incorrect | 7% | 8% | | 5% | 8% | | 5% | 10% | |
| Correct | 93% | 92% | | 95% | 92% | | 95% | 90% | |

p-values <0.01 were considered statistically significant and are bolded to indicate this.
Abbreviation: AD, Alzheimer's disease; Anxiety T-scores are scaled 0-100; APOE, apolipoprotein E; CFI, Cognitive Function Instrument; DSD, disease-specific distress; IGT-AD, Impact of Disclosure – Impact of Genetic Testing – AD; PHQ-9, Patient Health Questionnaire 9-items; PWB, Psychological Well Being Scale; Risk refers to participant self-perceived risk of developing AD; Relative risk means self-perceived risk of developing AD relative to others; T0, Baseline; T4, 1-7 days post pTau-217 disclosure; T5, 4-weeks post pTau-217 disclosure.
